# Understanding how demographic characteristics impact the level of physical activity children with neuromotor impairments experience while using a robot-assisted walker

**DOI:** 10.64898/2026.08.21.26361070

**Authors:** Jessica Youngblood, Markus Zaplachinski, Hua Shen, Elizabeth Condliffe

## Abstract

**Importance:** There are very few interventions designed for individuals with the most severe mobility impairments. Robotic walking may be an effective way to facilitate exercise in this population.

**Objective:** To examine how robot-assisted walker’s physical parameters and user characteristics moderate the exercise intensity achieved by individuals with neuromotor disorders causing mobility impairments.

**Design:** A prospective study.

**Intervention:** A single-session intervention involving an overground robot-assisted walker that can be used in an “endurance” mode requiring no voluntary movement or a “strength” mode during which voluntary movement could impact the gait pattern.

**Participants:** Individuals with pediatric-onset mobility impairments

**Main Outcome Measures:** Participants were characterized based on their age, sex, diagnosis, and Gilette Functional Assessment Questionnaire (FAQ) levels. Heart rate during the final minute of four 5-minute walking conditions: “strength” mode at fast speed, “strength” mode at slow speed, “endurance” mode at fast speed and “endurance” mode at slow speed was expressed as a percentage of each participant’s heart rate reserve (%HRR). Linear mixed-effects models were used to evaluate the impact of speed, device mode and user characteristics on the level of exercise achieved.

**Results:** 29 individuals (aged 2-26 years) with mobility impairments (FAQ levels 1-6) completed this study. Fast speeds were associated with a higher %HRR (β= 2.11, SE = 1.03, p = 0.044). Participants in FAQ class 1 exhibited significantly higher %HRR compared with those in FAQ classes 2 and 3 (β =18.6, SE=7.31, p=0.017; β= 16.9, SE = 8.13, p = 0.047, respectively). No other device or participant characteristics were associated with exercise intensity.

**Conclusions:** To facilitate higher exercise levels, users of robot-assisted walkers can increase their speed. Individuals who cannot take steps due to their neuromotor impairments experience the highest levels of exercise.

**Relevance:** The findings in this study highlight the promise of robot-assisted walkers to improve health, particularly in those who often face the greatest barriers to exercise.

## Introduction

People who cannot walk in community settings have difficulties exercising.^1–3^ Approximately one-third of the 3/1000 children in Canada with cerebral palsy (CP) cannot walk, and many other neuromuscular and rare genetic conditions can greatly impede ambulatory ability.^4,5^ Many people with CP and similar conditions live relatively sedentary lifestyles. For example, individuals who cannot walk unassisted due to CP spend ∼85-99% of their waking hours sedentary, compared to the average typically developing Canadian youth, who spends 8.5 hours (∼50%) sedentary.^3,6^ Inactivity is associated with the acquisition of comorbidities such as cardiovascular disease, hypertension and type 2 diabetes.^7^ Despite the prevalence and impacts of sedentariness, there are limited interventions to facilitate physical activity in this population, motivating research into novel therapeutic approaches.

The intensity of physical activity achieved during exercise can be measured as a percentage of heart rate reserve (%HRR), which is based on the heart rate attained by an individual during exercise relative to their resting and maximal heart rates.^8^ The spectrum of physical activity can be subcategorized into sedentary activity, any activity provoking less than 20%HRR, light physical activity (20-39%HRR), moderate (40-69%HRR) and vigorous physical activity (70-100%HRR)^9,10^. Among individuals who are largely sedentary, achieving even light physical activity can decrease the relative risk of secondary health conditions, and to an extent, more intense physical activity is even more effective.^11,12^

Robot-assisted walking is an emerging technology that shows promise in facilitating physical activity in populations who cannot walk. In addition to the more commonly studied impacts on motor function, robot-assisted walking appears to reduce consequences frequently associated with inactivity such as sleep disturbances, bowel function, and knee flexor spasticity.^13–15^ We recently confirmed that it can facilitate exercise in a population with severe mobility impairments.^16^ However, how to maximize the intensity of exercise and resultant health benefits is unknown. We believe the physical parameters of robot-assisted walkers, such as walking speed and degree of robot assistance, can be adjusted to optimize physical activity, and prescribing these parameters during clinical robot-assisted exercise sessions could assist in maximizing therapeutic benefits.^14^ The purpose of this study is to determine which robot-assisted walking parameters create higher exercise intensities based on individual characteristics. Therefore, the objectives of this study are to examine how walking speed and the level of robotic assistance impact the intensity of physical activity, and to investigate how user characteristics such as age, sex, diagnosis and functional ability moderate the intensity of exercise in conjunction with physical robot-assisted walking parameters.

## Methods

### Participants

This study was approved by the University of Calgary Conjoint Health and Research Ethics Board (REB21-1312).

Participants who were unable to walk independently in a community setting due to neuromotor impairment were recruited through convenience sampling. Participants were contacted if they had previously participated in research within the department of Clinical Neurosciences at the Alberta Children’s Hospital or had expressed interest in participating. Participants were included if they fit within the size constraints of an available Trexo Plus (Trexo Robotics, Mississauga, ON) robot-assisted walker, hereby referred to as the “Trexo” (femurthigh length >39m, calf length >50cm and weight <150 lbs). There were no limits on age or underlying diagnosis for this study, as the goal was to include the population that is interested in and able to use robot-assisted walkers to increase generalizability. Participants were excluded if they were unable to communicate pain or discomfort, were taking medications affecting heart rate, or had medical constraints to weight-bearing or aerobic activity (e.g. recent surgery).

### Device Description

Robot-assisted walking was facilitated using the Trexo. This device consists of a walker (Rifton Equipment, Rifton NY, United States), including a chest strap and an optional seat, supplemented with Trexo motors positioned laterally to the user’s hips and knees to assist joint movement during walking. This device can be adapted to an individual’s needs; it allows the option to remove the seat, and participants may face anteriorly or posteriorly, depending on each user’s trunk control._Individualized parameters, including range of motion, motor joint limits, mode (“strength” and “endurance”) and speed (ranging from 10-70 steps per minute) were set via a tablet interface connected through WiFi.

This robot–assisted walker has two modes that control the level of assistance provided while using the robot-assisted walker during walking. In “endurance” mode, the motors enforce the programmed gait cycle, regardless of the individual’s voluntary movement. In “strength” mode, wider bounds of tolerance allow greater user deviation from the target trajectory before assistance is provided. A caregiver or member of the research team were required to steer the robot-assisted walker during walking sessions.

### Study Design and Procedure

This was a prospective study involving single-session interventions to understand how different robot-assisted walking parameters impact physical activity. Prior to data collection, participants and/or the guardians provided written confirmation of consent and assent (when possible). Participants took part in two robot-assisted walking sessions: a familiarity session and a data collection session. The familiarity session was conducted to determine the personalized settings of the robot-assisted walker, including the participant’s preferred walking speed and to ensure the participant was comfortable with the device and our setting to reduce the impact of excitement or anxiety on their heart rate. Participants for whom the ideal walker settings were unclear were invited to attend a second session with another configuration.

At least 24 hours following their familiarity session, participants returned for a data collection session. At the start of the session, demographics (diagnosis, sex, age, and Gilette Functional Assessment Questionnaire walking scale (FAQ) levels) were collected from the parent or caregiver present. The FAQ is a parent-reported ordinal scale that provides insight into an individual’s functional ability, with level 1 (“cannot take any steps at all”) to level 10 (“walks, runs, and climbs on level and uneven terrain without difficulty or assistance”).^17^ Participants were then fitted with a Polar H9 heart rate monitor (®Polar Electro, Finland), which was synchronized with the smartphone application Elite HRV (Elite HRV, Gloucester, MA, United States of America) to record the beat-by-beat heart rate. At least 5-minutes of their resting heart rate was recorded, in which participants were instructed to relax in a seated or lying position based on their comfort. Caregivers often employed their own calming strategies (i.e. music, television); otherwise, a standardized calming video was played for the duration of the resting period. If more than six months had elapsed since initial fitting, participant leg lengths were re-measured before setup. Once comfortably positioned in the device, robot-assisted walking consisted of four 5-minute walking conditions: “strength” mode at fast speed, “strength” mode at slow speed, “endurance” mode at fast speed and “endurance” mode at slow speed. Slow and fast speeds were defined as 10 steps per minute below and above a participant’s determined comfortable walking speed. Conditions were presented in a randomized order with orders balanced across the group. Those unable to complete all four conditions in a single visit due to fatigue returned to complete those remaining at a later date. Heart rate was continuously recorded throughout the entirety of the robot-assisted walking session.

### Data Extraction

Exercise intensity during each robot-assisted walking condition was characterized by participants’ heart rate expressed as a percentage of their heart rate reserve (%HRR) during the final minute of walking to ensure that the participant’s heart rate had time to adapt to the activity intensity.^18^

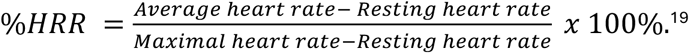

The average heart rate represents the instantaneous heart rate averaged across the final minute in each condition. The resting heart rate was defined as the lowest one-minute average calculated with a moving window average across the entire resting period. Maximal heart rate was predicted using the formula:

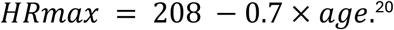

Some participants appeared to have excessive noise and were excluded. These were visually evident and confirmed by verifying that more than 1.75x the interquartile range above the 75^th^ percentile or below the 25^th^ percentile exceeded 10% of any analyzed minutes.

### Statistical Analysis

Level of physical activity in participants’ best conditions was analyzed to examine the effectiveness of robot-assisted walking at facilitating intense exercise in this group. All analyses were conducted in R (Posit, Boston, MA, United States of America). %HRR was analyzed using linear mixed-effects models to account for repeated measurements across walking conditions within participants. A random intercept for participants was included in all models, which was supported by substantial between-participant variability in baseline %HRR. Fixed effects were selected based on study design and clinical relevance, with an initial comprehensive model including fixed effects for walking speed and walking mode, and their interaction. Additional covariates (age, sex, diagnosis, FAQ class) were evaluated using a backward selection strategy guided by likelihood ratio tests and information criteria, while maintaining model parsimony. Interaction terms were examined but retained only if supported by both statistical evidence and adequate data support. Walking speed was specified a priori as the primary exposure of interest. Final model selection was based on nested model comparisons using maximum likelihood estimation, with statistical significance assessed at the 0.05 level.

The final model retained walking speed (fast vs slow) and FAQ class as fixed effects, with a random intercept for participant. Participants in FAQ levels 3 and above were classified into a single category (hereby referred to as FAQ class 3) to account for limited participants classifying as greater than level 3 and provide better fit to the data. Non-contributory covariates (age, sex, diagnosis) were removed based on lack of statistical support and negligible impact on key effect estimates. The walking mode main effect was also excluded after model comparison indicated no improvement in fit when retained.

Estimated marginal means and 95% confidence intervals were computed for walking speed and FAQ class to aid interpretation. Model assumptions were evaluated using residual diagnostics and quantile-quantile plots. Statistical significance of fixed effects was also assessed using Type III Wald F-tests with Satterthwaites approximation for degrees of freedom given the small sample size. Results are shown from all included participants’ data collection sessions, and descriptive statistics are reported as median (25^th^-75^th^ percentile).

## Results

32 individuals (aged 2-26 years) with moderate-to-severe mobility impairments (FAQ levels 1-6) participated in this study, three of whom were excluded due to excessive signal noise. Of the 29 participants whose data were used, 3 participated twice using different robot-assisted walking configurations (e.g. with and without a seat). Participants underlying diagnoses were categorized as cerebral palsy, other brain-based neurodevelopmental disorders, spine-based disorders (e.g. myelomeningocele), acquired conditions (e.g. traumatic brain injury), and neuromuscular disorders, with the largest portion having cerebral palsy (n=11, 40%) (Table 1). Four participants were unable to complete all robot-assisted walking conditions within a single session due to fatigue or equipment malfunction (e.g. heart rate monitor moving out of place) and finished the remaining conditions in an additional session. Participants had a median comfortable walking speed of 50 steps/minute (40-55 steps/minute).

**Table 1.**
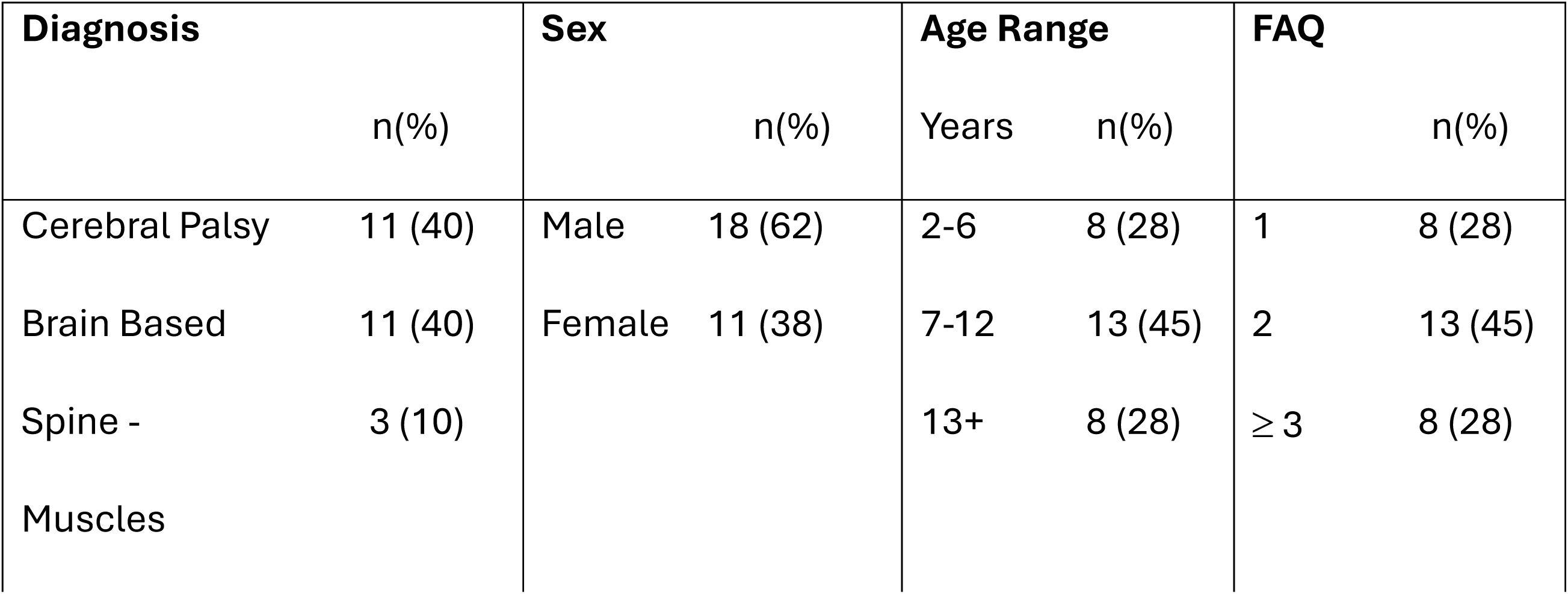
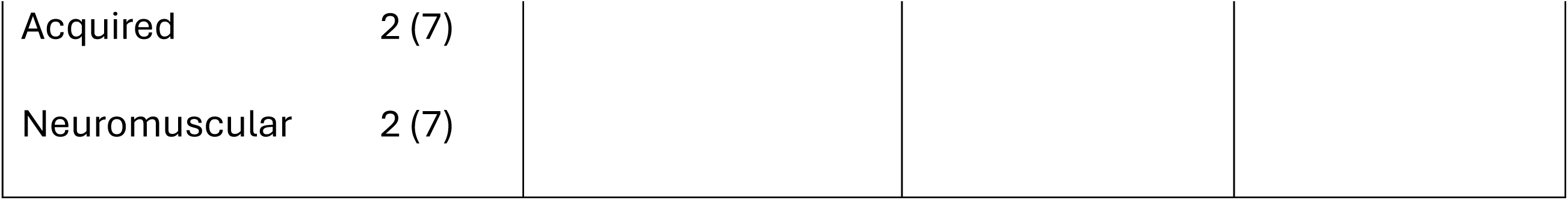
Participant demographics.

In “strength” mode, participants achieved 20.9%HRR (13.6-30.9) in their fast speed and 22.9%HRR (12.8-31.5) in their slow speed. In “endurance” mode, participants achieved 21.5%HRR (17.3-31.9) in their fast speed and 19.4%HRR (12.6-28.4) in their slow speed. Participants attained 28.5%HRR (20.6-35.1%HRR) in their most active condition, with 23/29 (79.3%) participants achieving at least light physical activity in their most active condition, and 13/29 (44.8%) achieving at least light physical activity in all conditions. 3/29 (10.3%) reached moderate physical activity in their best condition, and an additional 2/29 (6.9%) reached vigorous physical activity in their best condition.

In the final mixed-effects model, the inclusion of FAQ class significantly improved model fit compared with a model including speed alone (likelihood ratio test statistics 6.35, p = 0.042). Adjusting for functional ambulatory classification, walking speed was significantly associated with %HRR. Compared with walking in the slow conditions, fast walking was associated with a higher %HRR (beta = 2.11, SE = 1.03, p = 0.044). Functional ability, as measured by FAQ class, was also independently associated with %HRR. Participants in FAQ class 1 exhibited significantly higher %HRR compared with those in FAQ classed 2 and 3 (beta = 18.6, SE = 7.31, p = 0.017; beta = 16.9, SE = 8.13, p = 0.047, respectively).

The estimated marginal means analysis showed that %HRR was consistently higher during fast walking compared with slow walking for all FAQ classes. Specifically, in FAQ class 1, the adjusted mean %HRR increased from 37.7% (95% confidence interval: 25.3-50.1) at slow speed to 39.8% (27.4-52.2) at fast speed. Similarly, in FAQ class 2, mean %HRR increased from 19.1% (9.3-28.8) to 21.2% (11.4-30.9), and in FAQ class 3 from 20.8% (8.4-33.2) to 22.9% (10.5-35.3). Across all walking speeds, participants in FAQ class 1 exhibited substantially higher %HRR than those in FAQ classes 2 and 3, indicating increased exercise intensity among individuals with greater ambulatory impairment. The magnitude of the speed-related increase in %HRR was similar across FAQ classes, consistent with the absence of a statistically significant speed by FAQ interaction.

## Discussion

These results confirm that robot-assisted walking facilitates physical activity for children with severe neuromotor impairments and identify factors that impact exercise intensity. Increasing the speed of walking led to higher exercise intensities, while changing the mode of the robot-assisted walker did not consistently impact the intensity. When we examined how individual characteristics impact exercise intensity, we found that individuals with the most severe mobility impairments (FAQ 1) achieved higher exercise intensities while robot-assisted walking. However, age and diagnosis did not impact exercise intensity.

There are limited studies examining exercise intensity in children during robot-assisted walking. Our previous study found that all participants achieved at least light physical activity for at least a minute during a session of robot-assisted walking.^21^ In the current study, only 72% (23/32) of participants reached at least light physical activity in at least one condition, which is likely due to the study design. Here, the goal was to identify optimal settings for an individual, and exercise intensity was reported only at the 5^th^ minute of each condition. This constraint leads to under-representation of the exercise that can be achieved over an entire session. Studies of adults post-stroke and with spinal cord injuries in various devices show conflicting results, ranging from no physical activity achieved to reaching vigorous exercise intensity.^10,22,23^ Achieving any level of physical activity in children with severe mobility impairments is important, as they often spend the majority of their time sedentary, increasing their risk of developing multiple health conditions.^1^ There are currently no studies evaluating the specific characteristics that impact exercise intensity during robot-assisted walking. Understanding the effects of these characteristics could allow clinicians and parents to enhance the exercise intensity achieved during robot-assisted walking, which likely would decrease the participants’ risk of developing health conditions linked to a sedentary lifestyle.

The participants in this study achieved significantly higher %HRRs while walking at fast speeds compared to slow speeds. These results align with a study involving robot-assisted walking which found increased exercise in 5/5 participants with spinal cord injury when walking at their fastest comfortable speeds as opposed to their self-selected walking speeds.^23^ Similarly, in typically developing individuals, walking at “brisk” speeds creates light to moderate physical activity and leads to greater energy expenditure than walking at slow speeds.^24,25^ These findings indicate that robot-assisted walking users should walk as fast as can be comfortably tolerated if the goal is to maximize exercise.

Participants who are unable to take any steps (FAQ 1) experienced greater exercise intensity while robot-assisted walking than those with more walking ability. Individuals who are FAQ 1 experienced an estimated marginal mean exercise intensity just below the threshold for moderate physical activity, whereas children FAQ 2 C ≥3 did not or just barely achieved light physical activity. Since individuals FAQ 2 and ≥3 are more used to walking, they may be physiologically better-adjusted to exercise conditions, prompting less heart rate increase during robot-assisted walking.^26^ Further, robot-assisted walking enables walking and facilitates physical activity in individuals who are FAQ 1, which is important for decreasing health risks associated with a sedentary lifestyle.^27,28,1^

This study had several limitations. Heart rate data was only analyzed over one minute for each condition, which may not be representative of longer walking experiences. Familiarity sessions were implemented in an attempt to reduce the nervousness or excitement associated with participating in robot-assisted walking for the first time.

However, external factors may still have impacted heart rate during data collection sessions. Some participants struggled to fully rest during the resting heart rate data collection period, which may have caused inaccuracies in resting heart rate and, consequently, %HRR calculations. As a result, we may have underestimated the exercise intensities experienced by participants. The relatively small sample size and high prevalence of participants who were FAQ 1-3 limits generalizability.

## Conclusion

Robot-assisted walking facilitated physical activity for individuals with mobility impairments, especially for individuals with the most limited walking ability. Further, walking at fast speeds also increases the exercise intensity achieved. Clinicians and parents should be aware of this and encourage participants to walk as fast as they can. As mode did not impact exercise intensity, the mode used in robot-assisted walking sessions should be the one the child is the most comfortable in. Robot-assisted walking facilitating higher physical activity for those who are FAQ 1 is important as this group, that cannot take any steps, often faces the greatest challenges exercising. This study was the first to our knowledge to investigate how robot-assisted walker parameters and user characteristics influence exercise in individuals with severe neuromotor impairments. Future investigations with larger sample sizes are needed to understand if any additional user or device characteristics impact exercise intensity while robot-assisted walking.

## Data Availability

Most of the data that support the findings of this study are available openly, some data is held under restriction to protect participant privacy. Access to the open data and instructions for accessing the data are available on the "Understanding how demographic characteristics impact the level of physical activity children with neuromotor impairments experience while using a robot-assisted walker", https://doi.org/10.5683/SP4/FD9UUG, Borealis.

## Acknowledgments

We would like to thank our participants and the patient partners who guided and informed this study.

---

**Figure 1.**
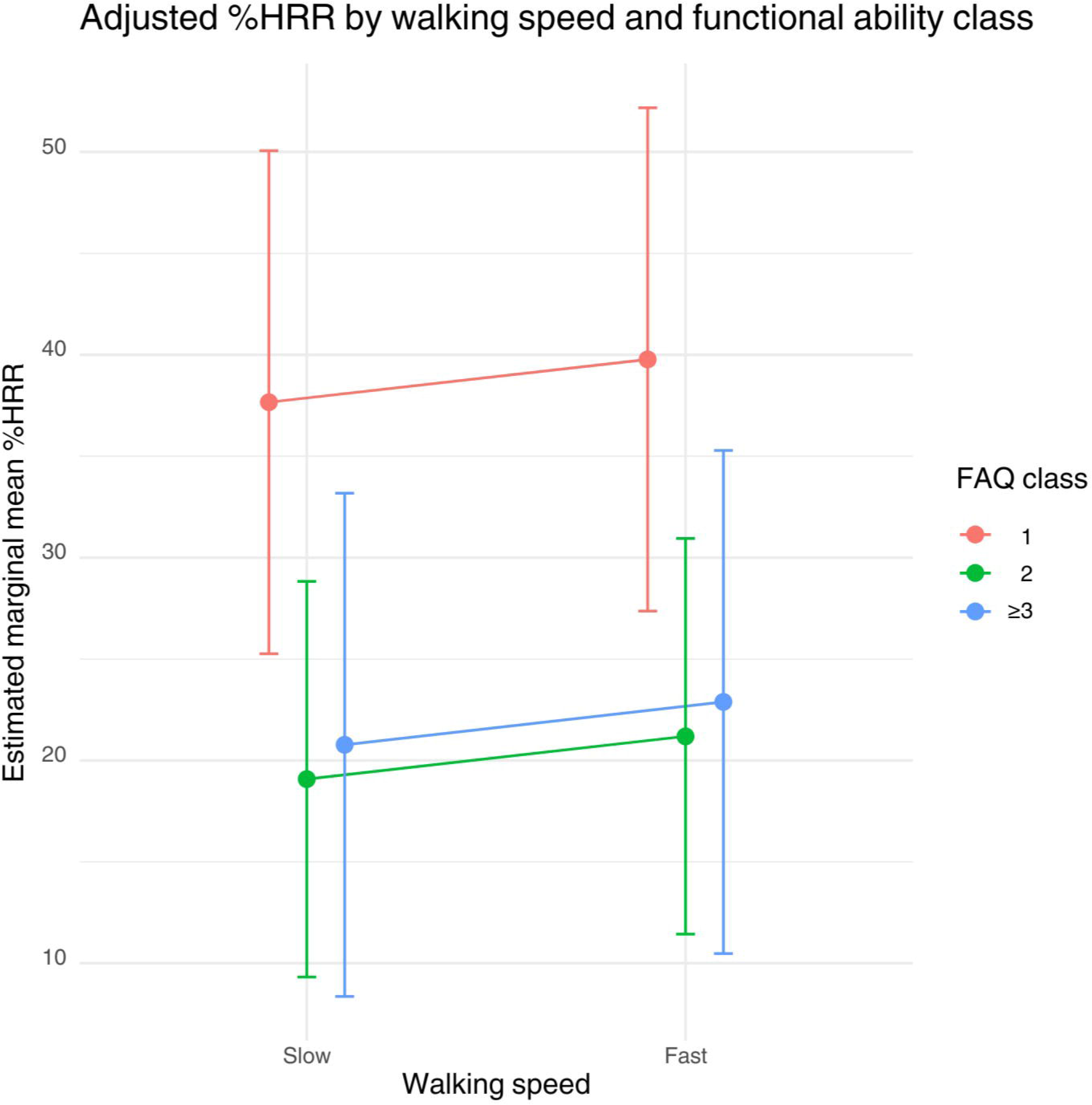
%HRR achieved during robot-assisted walking at slow and fast speeds. Results are shown as estimated marginal mean %HRR (95% confidence interval) for all FAQ classes.

